# Sleep quality worsened in an evening chronotype compared to others: a year-long longitudinal cohort study with prolonged lockdowns and restriction relaxations in RECETOX MU university employees

**DOI:** 10.1101/2023.09.22.23295980

**Authors:** Lucie Ráčková, Daniela Kuruczová, Irena Štěpaníková, Julie Bienertová-Vašků

## Abstract

**Background:** In 2020, people around the world were challenged by the outbreak of the COVID-19 pandemic. Countries responded with various restrictions, including lockdowns and stay-at-home orders, in an attempt to prevent the spread of the disease. Citizens were thus subjected to unprecedented uncertainty and stress. Prolonged exposure to such conditions may impact human health and well-being. One of the core aspects of proper physiological functioning is sleep. This prospective longitudinal study aims to investigate sleep quality and its relationship to chronotype over a year-long period from September 2020.

**Methods and findings:** Our year-long longitudinal prospective study focused on an employee cohort (N=54) at the Research Centre for Toxic Compounds in the Environment (RECETOX) of Masaryk University in the Czech Republic. During the first half of this period, three lockdowns with a cumulative duration of 100 days were imposed. During the second half of this period, the imposed restrictions were relaxed. Individuals were measured quarterly, i.e. at five time points. Sleep quality was measured using the Pittsburgh Sleep Quality Index (PSQI) while chronotype was established using the Reduced Morningness-Eveningness Questionnaire (rMEQ). We also used Perceived Stress Scale (PSS-14), Beck Depression Inventory (BDI-II), and General Anxiety Disorder-7 (GAD-7) to address potential confounders. The response rates of valid measurements across time points ranged from 87.04 % to 61.11 %. Our results show that sleep quality significantly worsened across the year for the evening chronotype but improved for the neutral and early chronotypes. Overall, over the year the incidence of poor sleep decreased by 16.13 % with 95% CI [-6.10%; 37.16%]. We did not find any significant sex differences in sleep quality. Perceived stress, symptoms of anxiety and depression were positively significantly associated with sleep problems in all measurements except in June. This study is limited by the small sample, decreasing number of individuals in chronotype categories and the lack of information on napping behavior.

**Conclusion:** These findings shed new light on the long-term influence of pandemic-related restrictions on individuals and particularly on the potentially more vulnerable evening chronotypes.

## Introduction

The pandemic outbreak of the coronavirus disease 2019 (COVID-19) resulted in an unprecedented challenge for societies around the world. The stressful atmosphere of uncertainty and restrictive governmental measures alone had influenced the psychophysiological functioning of the general population, as evidenced by a large number of publications [1–5]. One essential psychophysiological process which ensures overall health and regulates many functions of the organism is sleep [6]. The quality of sleep can be negatively affected by stress, resulting in sleep dysfunctions and the subsequent worsening of physical, psychological and cognitive functioning or increasing levels of perceived stress [7]. Therefore, understanding the influence of pandemics and related events on sleep quality in the general population is a crucial topic within the context of public health in general.

Existing research recognizes an association between pandemics and sleep problems. Several meta-analyses and systematic reviews summarized the evidence and found that the negative effect of COVID-19-related lockdowns on sleep quality was significant [8–12], but not uniform. Some individuals may have experienced sleep quality improvement [8,9]. This disparity might be caused by various factors. For example, social jetlag reduction and relaxation of work-related sleep restrictions [8], individual characteristics such as being an early chronotype [8], psychological factors, perception of stress, lifestyle factors [8–11], or geographical and socioeconomic factors [8,11]. The impact of sex and age seems to be complicated. Some studies found that younger individuals have better sleep quality during lockdown [8,9], but not all [8]. Regarding sex differences, studies suggest that women generally suffer from increased sleep [8,9], but, during confinement at home, sleep quality improved in women and worsened in men [12]. On the whole, differences in sleep quality changes during lockdown could be explained by a complex net of individual socio-psychological factors [8].

Most previously published studies were either cross-sectional [11,13,14] or short-term longitudinal [13,15–18]. Such designs are not necessarily well-suited to showing the long-term consequences of pandemics and related restrictions. Moreover, studies usually focused only on a single lockdown period [13,15–18]. Prospective studies which would include more than one lockdown period while also encompassing some time after the lockdown were lacking. Such studies would improve our understanding of the impact of repetitive measures employed by governments.

As the lockdown imposed on the inhabitants of the Czech Republic was not a continuous event, this study focuses on a year-long period from 1^st^ September 2020 to 31^st^ October 2021 in order to capture a representative image of both lockdown and post-lockdown phases. The epidemic trend of COVID-19 is shown in Fig. 1. From October 2020 to April 2021, three waves of COVID-19 prevalence emerged. The highest points were on 27^th^ October (1,200.01 cases per million), 10^th^ January 2021 (1,207.96 cases per million), and 5^th^ March 2021 (1,148.53 cases per million). Restrictions on free movement were employed from the 21^st^ October 2020 to the 12^th^ of April 2021 with stay-at-home policy phases from the 21^st^ of October 2020 to the 3^rd^ of November 2020, the 23^rd^ of December 2020 to the 14^th^ of February 2021, and 8^th^ March 2021 to 11^th^ April 2021. Data were obtained from Our world in data [19].

**Figure 1.:**
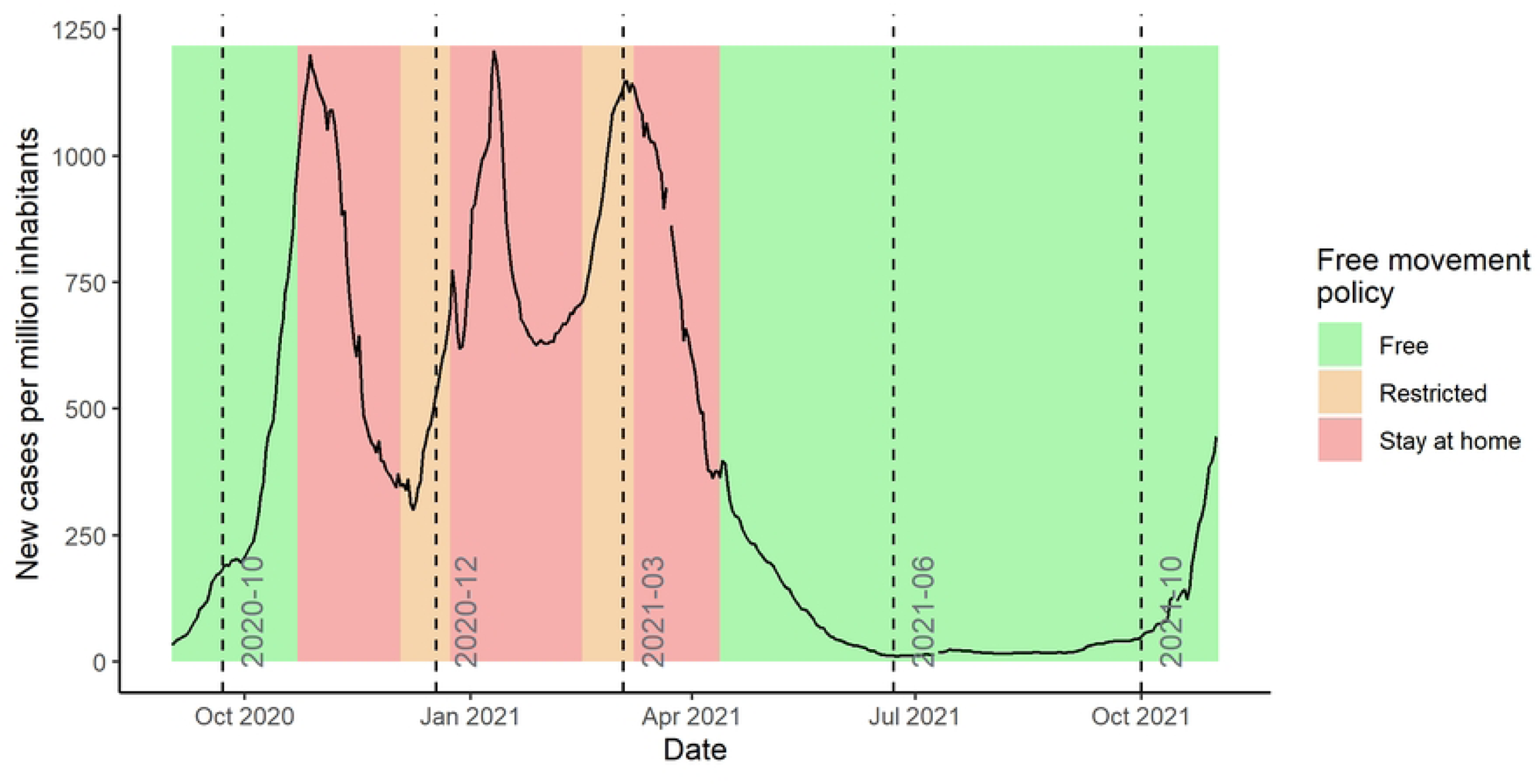
The national pandemic trend of COVID-19 prevalence and lockdown measures in Czech Republic from 1^st^ September 2020 to 31^st^ October 2021. The values are a 7-day rolling average of new cases per million inhabitants. Dashed lines represent the sending of the questionnaires.

This study aims to investigate the course of sleep quality and its relationship with chronotype. We used prospective longitudinal design in a year-long study which included three lockdown periods during the course of six months, and an equally long period of relaxed restrictions. We also examined cofactors such as perceived stress, and symptoms of depression or anxiety.

## Methods

### Subjects and study design

The study was approved by the Masaryk University (MU) Ethics Committee. The cohort consists of volunteers from the employees of the Research Center for Toxic Compounds in the Environment (RECETOX) at MU. All RECETOX employees were invited to the study via email in September 2020, before the restriction measures were tightened. The informed consent document was delivered by e-mail or in person. 56 people signed the informed consent document (either physically or electronically), and joined the study. Some characteristics of the population are in supplementary material S1 Table. We sent an e-mail to those who joined the study with links for questionnaires at the beginning of the study (October 2020) and then quarterly (December 2020, March 2021, June 2021, and September 2021), resulting in 5 time points of data collection. Some of these time points correspond to the beginning (October 2020, March 2021) and the end (December 2020, April 2021) of the strictest governmental restrictions in the Czech Republic. The course of national pandemic trends of COVID-19 disease and the timing of lockdown measures are in Fig. 1. The questionnaires were administered via Microsoft Office Forms (https://forms.office.com/).

### Measurements

#### Demographic data and COVID-19 positivity

We collected information on sex, age, nationality, number of children, and work position. In September 2020, December 2020, and April 2021, we asked participants if they had been diagnosed or were suspected of undergoing COVID-19 disease.

#### Chronotype

The morningness and eveningness of individuals were assessed using the Czech and English versions of the Reduced Morningness-Eveningness Questionnaire (rMEQ). It is a shorter 5-item version of the widely used Morningness-Eveningness Questionnaire. Cut-off scores are arbitrary with score ranges of 4–11 for the evening type, 12–17 for the neutral type, and 18–25 for the morning type. Evening types stay up later in the night and wake up later in the morning, morning types, on the other hand, go to sleep earlier in the evening and get up early in the morning. Psychometric parameters of the Czech rMEQ version were reported by Skočovský [20]. The appropriateness of arbitrary cut-off scores regarding the distribution of chronotypes in the population is discussed in several other studies [21,22].

#### Sleep quality

To evaluate sleep quality, we used the Czech and English versions of the Pittsburgh Sleep Quality Index (PSQI). The instrument was developed by Buysse et al. [23] and is currently the most commonly used measure in clinical and research settings [24]. It consists of 19 self-rated questions from which 7 components (subjective sleep quality, sleep latency, sleep duration, habitual sleep efficiency, sleep disturbances, use of sleep medication, daytime dysfunction) and global PSQI score can be calculated. PSQI also discriminates between good and poor sleepers using a cut-off score >5 of the global PSQI score [25]. The psychometric parameters of the Czech PSQI in a healthy population were recently reported [26].

#### Cofactors on emotional level

The RESTRESS study investigated several factors that may interfere with the quality of sleep, namely perceived stress, symptoms of depression, and anxiety.

Perceived stress was measured using the Czech and English versions of the Perceived Stress Scale (PSS-14). The scale consists of 14 items and was developed by Cohen et al. [27] for assessment of perceived unpredictability, uncontrollability and overload of respondents. The psychometric parameters of the English [28] and Czech version were previously reported [29].

Symptoms of depression were measured using the Czech and English versions of the Beck Depression Inventory (BDI-II). This 21-item inventory was developed by Beck et al. [30] and shows good psychometric parameters [31,32].

General anxiety symptoms were evaluated by the 7-item Generalized Anxiety Scale. Spitzer et al. [33] for brief clinical measurement of anxiety disorder. The psychometric parameters were previously tested in another studies [34,35].

#### Statistical analysis

Subjects with complete data from the demographic questionnaire and at least one data point in rMEQ and PSQI were included in the analysis. Data analysis was performed using statistical software R version 4.1.2 [36]. As a first step of the analysis, descriptive statistics for all variables across all time points were calculated and basic relationships between variables were explored. As part of the exploratory analysis, subjects with complete and incomplete data across time points were compared to identify potential differences. R library rmcorr [37] was used to determine the repeated measures correlation of variables over time. Next, R package lmerTest [38] was used for repeated measures ANOVA and package nlme [39] for follow-up comparisons. For both rMEQ and PSQI, basic psychometric properties and correlations across time were assessed to ensure their reliability. Effect sizes were interpreted using Gignac and Szodorai [40] guideline. Dominant chronotype was calculated for all subjects using mean rMEQ score throughout all time points and chronotype was used in the further analysis as a time-stable variable. To test the relationship between sleep quality and chronotype across time, a linear mixed effects model (LMM) with random intercept was fitted using lmerTest R library [38]. The dependent variable was set to be the PSQI score, with time point, rMEQ category, and their interaction as covariates.

To evaluate the representativeness of our sample, a basic comparison was performed with data obtained from the RECETOX human resources department. Expected and observed frequencies for sex, age group, and job category were compared using Pearson’s chi-squared test.

## Results

### Participants

The analyzed sample consisted of 54 individuals (29 female) with a mean age of 37.6 (*SD* = 8.05) years who responded in at least one time point. The majority of the sample were Czechs (34 individuals, 62.96 %), 4 individuals were Slovaks (7.41 %), and 11 individuals were from 9 other countries (details in S2 Table). There were 29 participants (53.70 %) with at least one child (details in S2 Table). 25 individuals are complete cases that responded to every time point, and the other 29 individuals responded to at least one questionnaire. The response rates are in supplementary material S3 Table. Complete and incomplete cases did not differ in demographic parameters (details in S4 Table). Compared to the entire population of RECETOX employees, our sample has similar distribution of sex categories, age categories, and research positions (23 individuals, 47.92 %). On the other hand, academic positions (14 individuals, 29.17 %) are overrepresented and administrative positions (11 participants, 22.92 %) are underrepresented (details in S1 Table).

The predominant chronotype was the neutral chronotype (27 individuals, 50.00 %), the second was the morning chronotype (19 individuals, 35.19 %), and the least represented was the evening chronotype (8 individuals, 14.81 %; details in S5 Table). Individual changes in the chronotype category are visualized in Fig. 2. At the beginning of the study, 55.32 % of respondents had poor sleep according to the PSQI criteria. The number of poor sleepers was lowered through the study, resulting in 41.18 % of poor sleepers (Fig. 3; details in S5 Table). Based on McNemar’s test on paired proportions, the estimated difference between the proportion of people with poor sleep at the end of the study and at the beginning of the study is a reduction of 16.13%, 95%CI [-6.10%; 37.16%]. The chi-squared test did not show significant differences in sleep quality between the complete and missing cases (see S4 Table). The majority of the sample (41 individuals, 80.39 %) had not suspected nor confirmed SARS-CoV-2 infection during the study. Only 2 individuals (3.92 %) had confirmed, and 8 individuals (15.69 %) were suspected of SARS-CoV-2 infection during the study (see S7 Table).

**Figure 2.:**
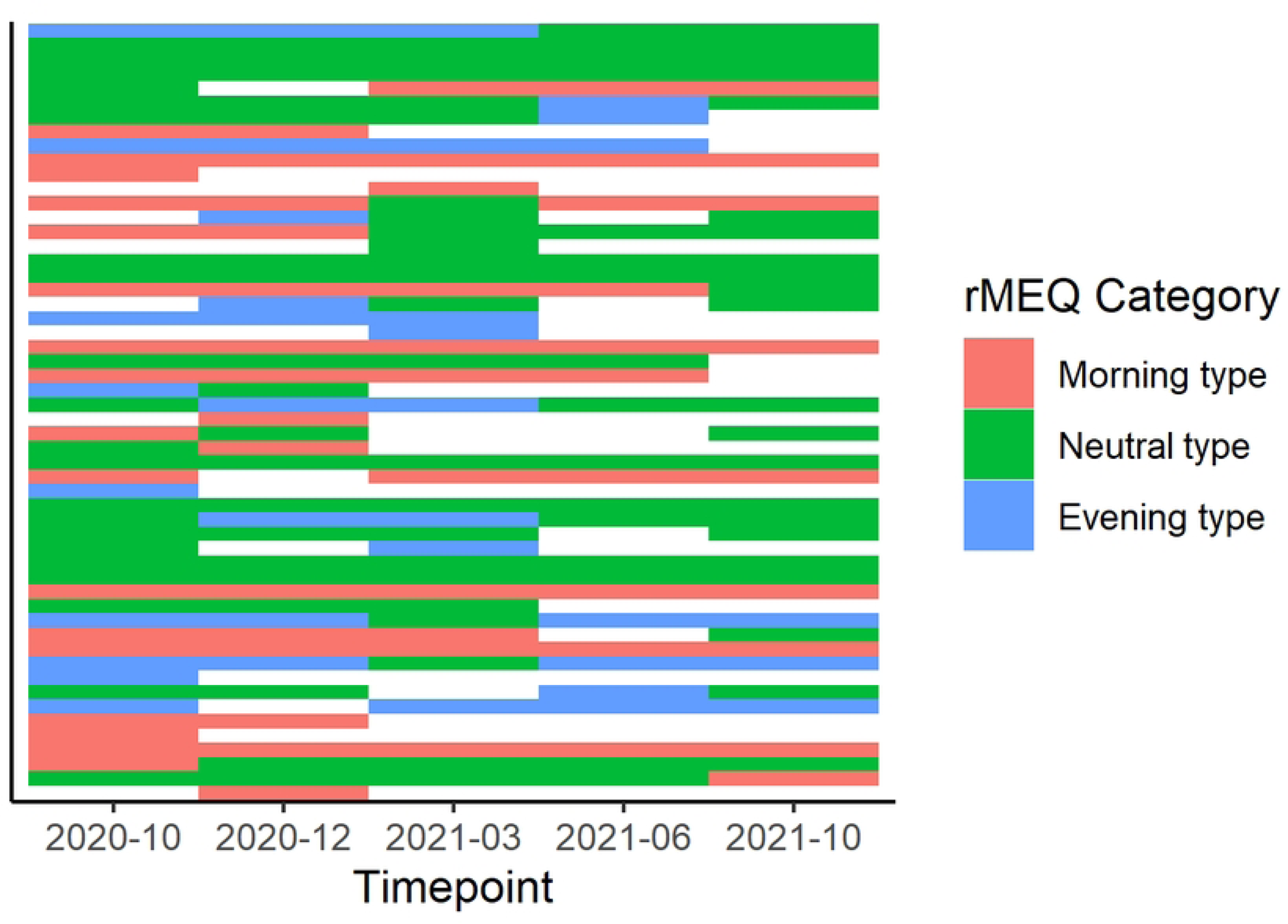
Individual changes in chronotype categories (measured by rMEQ) over the course of the study.

**Figure 3.:**
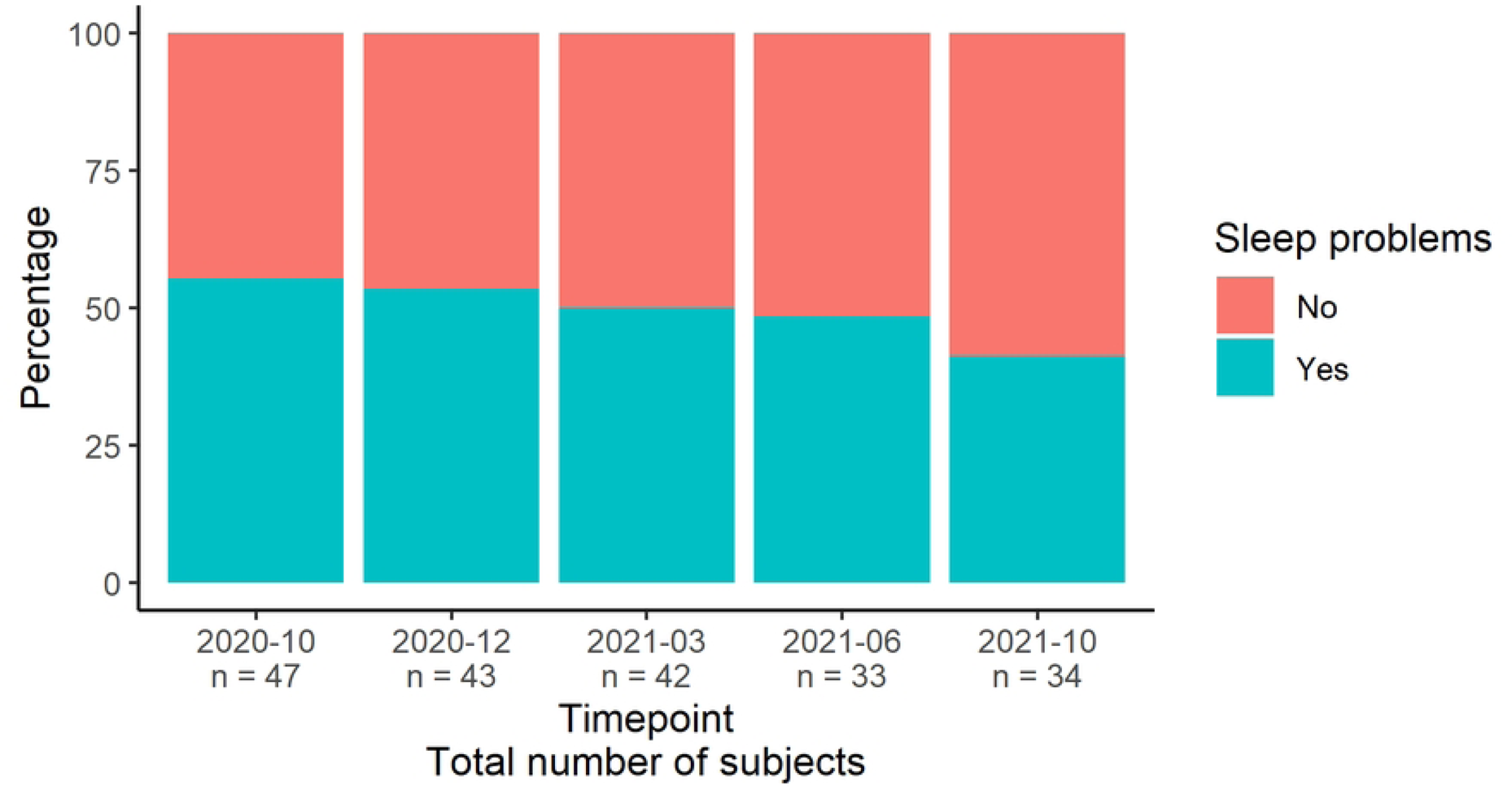
Good and poor sleepers’ ratio (according to PSQI criteria) over the course of the study.

Perceived stress was increasing from October (*M =* 24.50, *SD* = 7.04) to March 2021 (*M* = 27.53, SD = 9.05), followed by decrease in July 2021 (*M =* 21.74, *SD =* 8.92) and slight increase in October. Symptoms of anxiety had similar course. <u>Mean</u> values were increasing from October (*M* = 6.33, *SD* = 4.62) to March 2021 (*M* = 8.00, *SD* = 5.78) and then decreased in June (*M* = 4.65, *SD* = 4.33) and slightly increased in October 2021 (*M* = 5.03, *SD* = 4.45). Lastly, measurements of depression were somewhat similar. In October 2020, the mean BDI-II score was 8.85 (*SD* = 8.87), then was increasing with the highest mean score in March 2021 (*M* = 7.97, *SD* = 12.04). Contrary to PSS-14 and GAD-7, the lowest mean score of BDI-II was measured in October 2021 (*M* = 7.21, *SD* = 9.40). More details are in the S6 Table. Alcohol intake, sleeping pills consumption and cigarette smoking are reported in S11 Table.

### Exploratory analysis

To examine the relationship between sleep quality variables and chronotype, the repeated measures correlation was calculated (results in S8 Table). The increase in PSQI global score was significantly and weakly negatively correlated with the rMEQ score, *r* (144) = −.22, p = .009. Meaning that worse sleep quality correlated with evening chronotype tendency. Next, bedtime was significantly moderately negatively correlated (*r* (144) = −.28, p = .001), and sleep time weakly significantly positively correlated (*r* (144) = .23, p = .005) with the rMEQ score. Wake-up time was not significantly correlated with the rMEQ score. This indicates that the morning chronotypes slept longer than the evening chronotype.

Further analysis of sleep quality, chronotype, and demographic variables using ANOVA (S9 Table) revealed that wake-up time and bedtime were significantly associated with chronotype, complementing the results of the correlation analysis above. However, the ANOVA revealed no statistically significant difference in sleep quality between chronotype categories, or in sleep time between chronotype categories. Sex differences were not apparent in any variable.

Repeated measures correlation analysis with cofactors revealed significant moderate positive association of sleep quality and perceived stress, r (130) = .27, p=.002, and with symptoms of anxiety, r (128) = .20, p = 0.02. Correlation with depressive symptoms was positive and large, r (122) = .38, p < 0.001. Interestingly, these associations were not present in June 2021. In chronotypes, no statistically significant correlations were found from October 2020 to March 2021. However, in June 2021, we found significant large negative correlation with perceived stress, r = −.47, p = 0.03, and borderline significant large negative correlation with symptoms of depression, r = −.0.37, p = 0.09. In October 2021, there was borderline significant moderate negative correlation with perceived stress, r = −.29, p = .09. Correlation with depressive symptoms was significant and large, r = −.48, p = 0.005, and also with symptoms of anxiety, r= −.36, p = 0.04. More details are in S8 Table.

Similar results were obtained through mixed ANOVA (S9 Table), where higher values of perceived stress, symptoms of depression and anxiety associated with worse sleep quality. This trend was present in majority of timepoints, except for June 2021 where all went statistically insignificant. No significant association was found with chronotype with exception of June and October 2021 where borderline significant association was found with perceived stress.

### Change of sleep and chronotype over the time

To assess whether chronotype category and time predicted changes in sleep quality, we used LMM. Sleep quality was significantly predicted by time (see tab. 1). With increasing time, sleep quality improved. However, the opposite direction was found in the interaction between evening type and time (see also Fig. 4). Interaction between morning type and time was not statistically significant (more information in S10 Table)

**Figure 4.:**
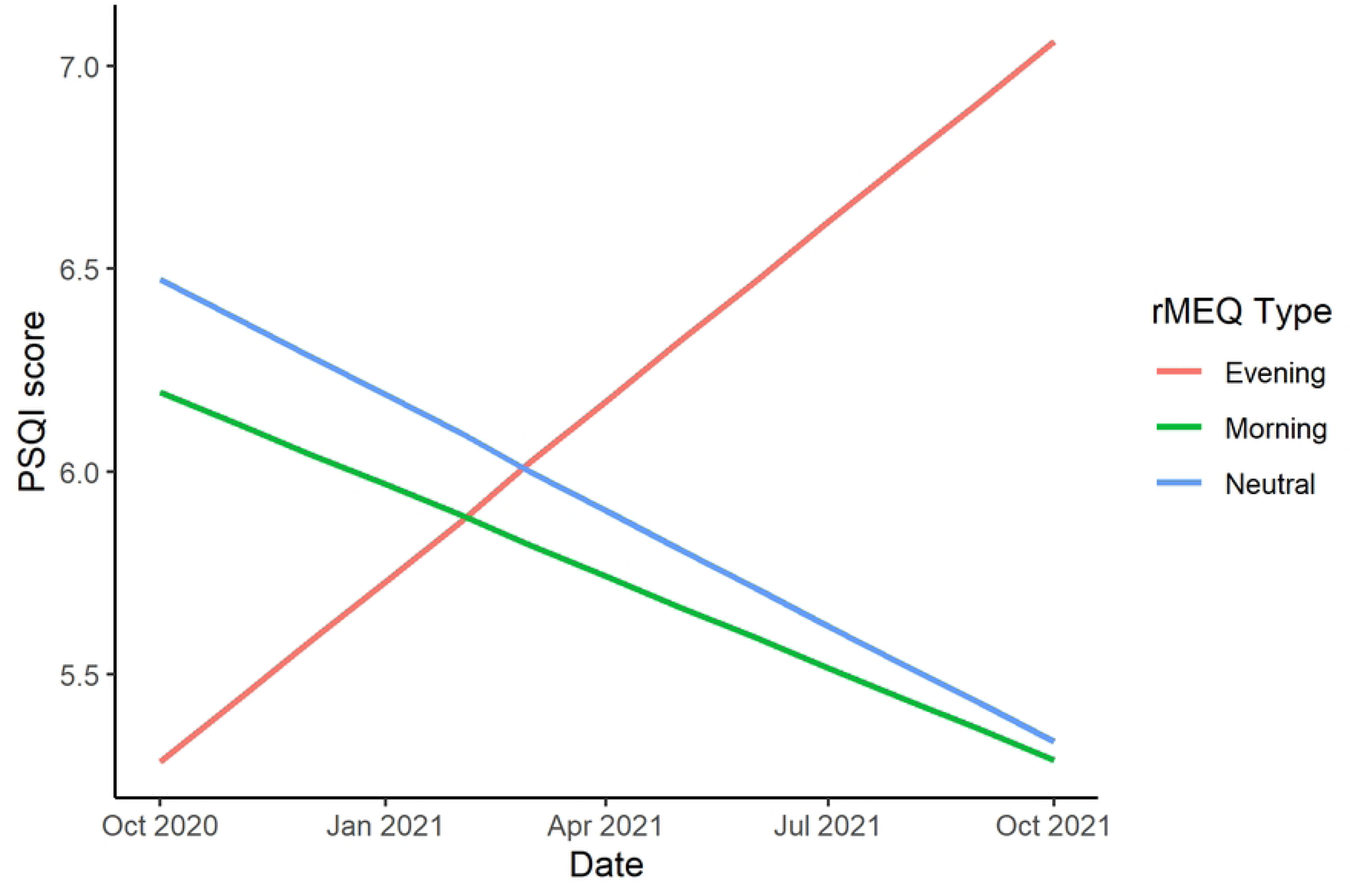
Graphical representation of the estimated relationship between time and sleep quality (PSQI Global score) in morning chronotype (green line), neutral chronotype (blue line), and evening chronotype (red line).

**Table 1.:** Results of the LMM model to test the effect of chronotype and time on sleep quality. Statistically significant predictors (p<0.05) are shown in bold.

| Variable |  | Estimate | SE | p-value |
| --- | --- | --- | --- | --- |
| Intercept |  | 6.474 | 0.500 | <2e-16 |
| rMEQ category | Neutral type | ref. | - | - |
|  | Evening type | -1.190 | 1.086 | 0.276 |
|  | Morning type | -0.280 | 0.782 | 0.722 |
| Time [months] |  | **-0.095** | **0.047** | **0.046** |
| Time x rMEQ cat. | Time x Neutral type | ref. | - | - |
|  | Time x Evening type | **0.243** | **0.121** | **0.047** |
|  | Time x Morning type | 0.020 | 0.078 | 0.803 |

## Discussion

To our knowledge, this is the first study that assesses the sleep problems and chronotype longitudinally and non-retrospectively, encompassing three government-proclaimed stay-at-home lockdown periods (with a cumulative length of 100 days) and an equally long period of relaxation of restrictions. The studied sample was drawn from the population of university workers at RECETOX, MU (Czech Republic). We did not find significant general differences in sleep quality between chronotypes. However, we found that evening chronotypes had better sleep quality at the beginning of the study, which continuously worsened during all lockdown periods and subsequent 6 months of restriction relaxation in the evening chronotype. This result is in line with findings from Bazzani et al. [41], but contradict with others that did not find differences in the course of sleep quality in different chronotypes with chronotype and sleep quality changes [18,42,43]. Surprisingly, contrary to other studies [11–13,44–49], we found no sex differences in sleep quality. Our cohort has a small number of respondents who underwent or were suspected of SARS-CoV-2 infection (3.92 % and 15.69 % respectively). We can therefore assume that the results of this study may indicate the impact of pandemic measures rather than the consequences of possible COVID-19 infection or related treatment.

Our results extend the previous findings on the greater vulnerability of evening chronotype to sleep problems during the lockdown period [13,41,50] and in general [51–53]. This is somewhat contrary to expectations, because more relaxed working hours during stay-at-home orders may have benefitted evening types with prolonged wake-up time and more sleep, as observed by Staller and Randler [54] Hence, it could be hypothesized that the tendency for evening chronotype could emphasize sleep problems not only during stressful life periods but with possible long-lasting effects.

Another important finding was that the ratio of poor sleepers in the sample declined by 16.13 % at the end of the study. This finding is contrary to previous studies which found an increased prevalence of sleep problems during lockdowns [11,13,47,48,55,56]. Based on their findings we would expect sleep problems to graduate during the lockdown and then mitigate through the relaxation of restrictions. The observed trend in our study may be caused by successful coping with the pandemic situation, or by individual differences in response to restrictive measures. Another explanation might be in the initial number of poor sleepers. Some studies reported [57,58] that individuals with present sleep problems may experience sleep improvement during the pandemic. Our sample had a higher ratio of poor sleepers than was found in several meta-analyses (prevalence in the general population was 27 % in Alimoradi et al. [12], 18% in Alimoradi et al. [11], 32.3 % in Jahrami et al. [59], 36.73 % in Jahrami et al. [60], but corresponds with a range of 17.65 – 81 % estimated by Lin et al. [8]). It may be that our participants with a higher prevalence of poor sleepers benefited from social and system changes during the pandemic by improving their sleep.

Another interesting associations were found in correlation analyses with potential cofactors, such as perceived stress, symptoms of depression and anxiety. During lock-down period, these cofactors were in statistically significant positive association with sleep quality but not with chronotype. However, different results were found in the period of restriction relaxation. In June 2021, there were no significant correlations with sleep quality, but significant large negative correlation between chronotype, depressive symptoms and perceived stress emerged. And in October 2021, the correlations were significant in all cofactors and for both sleep quality and chronotype. These chronotype specific changes are somewhat similar to findings from Salfi et al. [42] who in their longitudinal study found relationship between survey waves, evening chronotype and perceived stress, but not with symptoms of depression and anxiety. Such dynamics might be partially explained by seasonal effects [61–63]. Another possible explanation may be the psychosocial effect of COVID-19 related restriction policies changes. In our data, during the period of restriction relaxation, the symptoms of anxiety, depression and perception of stress decreased, while the mean sleep quality remained relatively stable. As a consequence, associations between evening chronotype and higher tendency for stress perception and depressive symptoms were highlighted in the data. It is also possible that pandemic had longer lasting impact on evening chronotypes further increasing negative impact on their mood. Apart from that, this outcome challenges suggestions of some studies which found that sleep quality is a predictor of mental health [64] with evidence for bi-directional relationship [65,66]. Further research should investigate the bi-directional relationship between mood and sleep quality in longitudinal cohorts to address strength of the relationship with respect to the impact of major life or societal events, with specific focus on the vulnerability of evening chronotype.

This study is limited by the small sample which may have reduced the power of the study and did not allow the inclusion of more relevant variables in the model (e.g., symptoms of depression, anxiety, or perceived stress). Moreover, the number of individuals in chronotype categories decreased with time, most prominently in the evening chronotype category. In terms of the generalizability of the results, our cohort overrepresented academics and underrepresented administrative positions. Our study also did not include information on naptimes, use of soft drinks, medication and drugs, or data on the period before the pandemic and the first half year of the pandemic. Despite its limitations, the value of this study lies in the combination of longitudinal study design and a unique, unprecedented situation. Its results certainly add to our understanding of the long-lasting influence of pandemics and related restrictions on sleep quality in different chronotypes.

Results from our study provide evidence for higher vulnerability of evening chronotypes to the negative effects of pandemics on sleep quality and moods with long lasting effects. Future research should use larger sample, and include other symptoms interfering with sleep quality, e.g. depression, anxiety, perceived stress, etc. Furthermore, studies need to be carried out to determine effective countermeasures for these negative consequences of major psychosocial life events in evening chronotypes. These may include cognitive behavioral therapy for insomnia [67,68], digital therapeutics, [67], or other recommendations (summarized in Altena et al. [68]). Findings from such studies may have important implications for developing prevention programs for the general public implemented by the government, public health professionals, and employers during possible future lockdowns. These programs may include educational projects on good sleep practices, and mental, emotional, and social health [67].

## Data Availability

o Because the study involves human research participants from a relatively small and closed group of participants, and includes their personal and sensitive data, we decided to provide the data set in anonymized form upon request. Requests for data can be sent to. The data underlying the results presented in the study are available in the Supplementary Material.

## Acknowledgments

Authors would like to thank RECETOX MU for their support and to the individuals who participated in the study. We would also like to thank Tomáš Piskovský for assistance with the data analysis and David Konečný for proofreading the manuscript.

## Supporting Information

**S1 Table. Characteristics of the studied population and study sample. S2 Table. Demographic parameters of the study sample.**

**S3 Table. Response rates for questionnaires in individual time points.**

**S4 Table. Demographic parameters and sleep problems in complete and incomplete cases.**

**S5 Table. Distribution of sleep quality and chronotype in different timepoints, sex and age groups.**

**S6 Table. Descriptive statistics of perceived stress, symptoms of anxiety and depression in different timepoints.**

**S7 Table. COVID-19 status of the studied population.**

**S8 Table. Repeated measures correlation. Statistically significant results are in bold.**

**S9 Table. ANOVA. Statistically significant results are in bold.**

**S10 Table. Linear mixed model fit by REML. t-tests use Satterthwaite’s method.**

**S11 Table. Alcohol intake, sleeping pills consumption, and cigarette smoking.**

## References

1. Nochaiwong S, Ruengorn C, Thavorn K, Hutton B, Awiphan R, Phosuya C, et al. Global prevalence of mental health issues among the general population during the coronavirus disease-2019 pandemic: a systematic review and meta-analysis. Sci Rep [Internet]. 2021 May 13;11(1):10173. Available from: 10.1038/s41598-021-89700-8

2. Salari N, Hosseinian-Far A, Jalali R, Vaisi-Raygani A, Rasoulpoor S, Mohammadi M, et al. Prevalence of stress, anxiety, depression among the general population during the COVID-19 pandemic: a systematic review and meta-analysis. Global Health [Internet]. 2020;16(1):57. Available from: 10.1186/s12992-020-00589-w

3. Richter D, Riedel-Heller S, Zürcher SJ. Mental health problems in the general population during and after the first lockdown phase due to the SARS-Cov-2 pandemic: rapid review of multi-wave studies. Epidemiol Psychiatr Sci [Internet]. 2021/03/09. 2021 Mar 9;30:e27. Available from: https://www.cambridge.org/core/article/mental-health-problems-in-the-general-population-during-and-after-the-first-lockdown-phase-due-to-the-sarscov2-pandemic-rapid-review-of-multiwave-studies/6186D1BEFB5515F92FF828CF84CAFB6D

4. Robinson E, Sutin AR, Daly M, Jones A. A systematic review and meta-analysis of longitudinal cohort studies comparing mental health before versus during the COVID-19 pandemic in 2020. J Affect Disord [Internet]. 2022;296:567–76. Available from: https://www.sciencedirect.com/science/article/pii/S0165032721010570

5. Dragioti E, Li H, Tsitsas G, Lee KH, Choi J, Kim J, et al. A large-scale meta-analytic atlas of mental health problems prevalence during the COVID-19 early pandemic. J Med Virol [Internet]. 2022 May 1;94(5):1935–49. Available from: 10.1002/jmv.27549

6. Zielinski MR, McKenna JT, McCarley RW. Functions and Mechanisms of Sleep. AIMS Neurosci. 2016;3(1):67–104.

7. Lo Martire V, Caruso D, Palagini L, Zoccoli G, Bastianini S. Stress & sleep: A relationship lasting a lifetime. Neurosci Biobehav Rev [Internet]. 2020;117:65–77. Available from: https://www.sciencedirect.com/science/article/pii/S0149763419301496

8. Lin YN, Liu ZR, Li SQ, Li CX, Zhang L, Li N, et al. Burden of Sleep Disturbance During COVID-19 Pandemic: A Systematic Review. Nat Sci Sleep [Internet]. 2021 Jun;Volume 13:933–66. Available from: https://www.dovepress.com/burden-of-sleep-disturbance-during-covid-19-pandemic-a-systematic-revi-peer-reviewed-fulltext-article-NSS

9. Pilcher JJ, Dorsey LL, Galloway SM, Erikson DN. Social Isolation and Sleep: Manifestation During COVID-19 Quarantines. Front Psychol [Internet]. 2022;12. Available from: https://www.frontiersin.org/articles/10.3389/fpsyg.2021.810763

10. Cipriani GE, Bartoli M, Amanzio M. Are Sleep Problems Related to Psychological Distress in Healthy Aging during the COVID-19 Pandemic? A Review. Int J Environ Res Public Health. 2021 Oct;18(20).

11. Alimoradi Z, Broström A, Tsang HWH, Griffiths MD, Haghayegh S, Ohayon MM, et al. Sleep problems during COVID-19 pandemic and its’ association to psychological distress: A systematic review and meta-analysis. EClinicalMedicine [Internet]. 2021 Jun;36:100916. Available from: https://linkinghub.elsevier.com/retrieve/pii/S2589537021001966

12. Alimoradi Z, Gozal D, Tsang HWH, Lin C, Broström A, Ohayon MM, et al. Gender-specific estimates of sleep problems during the COVID-19 pandemic: Systematic review and meta-analysis. J Sleep Res [Internet]. 2022 Feb 9;31(1). Available from: https://onlinelibrary.wiley.com/doi/10.1111/jsr.13432

13. Salfi F, Lauriola M, D’Atri A, Amicucci G, Viselli L, Tempesta D, et al. Demographic, psychological, chronobiological, and work-related predictors of sleep disturbances during the COVID-19 lockdown in Italy. Sci Rep [Internet]. 2021 Dec 1;11(1):11416. Available from: http://www.nature.com/articles/s41598-021-90993-y

14. Robillard R, Dion K, Pennestri M, Solomonova E, Lee E, Saad M, et al. Profiles of sleep changes during the COVID-19 pandemic: Demographic, behavioural and psychological factors. J Sleep Res [Internet]. 2021 Feb 17;30(1). Available from: https://onlinelibrary.wiley.com/doi/10.1111/jsr.13231

15. Zhang Y, Zhang H, Ma X, Di Q. Mental Health Problems during the COVID-19 Pandemics and the Mitigation Effects of Exercise: A Longitudinal Study of College Students in China. Vol. 17, International Journal of Environmental Research and Public Health. 2020.

16. Martínez-de-Quel Ó, Suárez-Iglesias D, López-Flores M, Pérez CA. Physical activity, dietary habits and sleep quality before and during COVID-19 lockdown: A longitudinal study. Appetite [Internet]. 2021;158:105019. Available from: https://www.sciencedirect.com/science/article/pii/S019566632031641X

17. Saraswathi I, Saikarthik J, Kumar KS, Srinivasan KM, Ardhanaari M, Gunapriya R. Impact of COVID-19 outbreak on the mental health status of undergraduate medical students in a COVID-19 treating medical college: a prospective longitudinal study. PeerJ. 2020;8:e10164.

18. Gao C, Scullin MK. Sleep health early in the coronavirus disease 2019 (COVID-19) outbreak in the United States: integrating longitudinal, cross-sectional, and retrospective recall data. Sleep Med [Internet]. 2020;73:1–10. Available from: https://www.sciencedirect.com/science/article/pii/S1389945720302999

19. Ritchie H, Mathieu E, Rodés-Guirao L, Appel C, Giattino C, Ortiz-Ospina E, et al. Coronavirus Pandemic (COVID-19) [Internet]. Our World in Data. 2022. Available from: https://ourworldindata.org/coronavirus

20. Skočovský KD. Psychometrické vlastnosti české verze Dotazníku ranních a večerních typů (MEQ). Sociální procesy a osobnost Brno Psychol ústav FF MU v Brně. 2003;260–7.

21. Caci H, Deschaux O, Adan A, Natale V. Comparing three morningness scales: Age and gender effects, structure and cut-off criteria. Sleep Med. 2009;10(2):240–5.

22. Loureiro F, Garcia-Marques T. Morning or Evening person? Which type are you? Self-assessment of chronotype. Pers Individ Dif [Internet]. 2015;86:168–71. Available from: 10.1016/j.paid.2015.06.022

23. Buysse DJ, Reynolds CF, Monk TH, Berman SR, Kupfer DJ, Buysse DJ, Reynolds CF, Monk TH, Berman SR, Kupfer DJ. The Pittsburgh Sleep Quality Index: a new instrument for psychiatric practice and research. Psychiatry Res. 1989;28:193–213. 1989;

24. Mollayeva T, Thurairajah P, Burton K, Mollayeva S, Shapiro CM, Colantonio A. The Pittsburgh sleep quality index as a screening tool for sleep dysfunction in clinical and non-clinical samples: A systematic review and meta-analysis. Sleep Med Rev [Internet]. 2016;25:52–73. Available from: 10.1016/j.smrv.2015.01.009

25. Buysse DJ, Reynolds CF 3rd, Monk TH, Berman SR, Kupfer DJ. The Pittsburgh Sleep Quality Index: a new instrument for psychiatric practice and research. Psychiatry Res. 1989 May;28(2):193–213.

26. Manková D, Dudysová D, Novák J, Fárková E, Janků K, Kliková M, et al. Reliability and Validity of the Czech Version of the Pittsburgh Sleep Quality Index in Patients with Sleep Disorders and Healthy Controls. Biomed Res Int. 2021;2021.

27. Cohen S, Kamarck T, Mermelstein R. A global measure of perceived stress. J Health Soc Behav. 1983;385–96.

28. Lee EH. Review of the Psychometric Evidence of the Perceived Stress Scale. Asian Nurs Res (Korean Soc Nurs Sci) [Internet]. 2012;6(4):121–7. Available from: https://www.sciencedirect.com/science/article/pii/S1976131712000527

29. Figalová N, Charvát M. The perceived stress scale: Reliability and validity study in the Czech Republic. Cesk Psychol [Internet]. 2021 Feb 28;65(1 SE-):46–59. Available from: https://ceskoslovenskapsychologie.cz/index.php/csps/article/view/33

30. Beck AT, Steer RA, Ball R, Ranieri WF. Comparison of Beck Depression Inventories-IA and-II in psychiatric outpatients. J Pers Assess. 1996;67(3):588–97.

31. Wang YP, Gorenstein C. Psychometric properties of the Beck Depression Inventory-II: a comprehensive review. Brazilian J Psychiatry. 2013;35:416–31.

32. Ociskova M, Prasko J, Kupka M, Marackova M, Latalova K, Cinculova A, et al. Psychometric evaluation of the Czech Beck Depression Inventory-II in a sample of depressed patients and healthy controls. Neuroendocr Lett. 2017;38(2):98– 106.

33. Spitzer RL, Kroenke K, Williams JBW, Löwe B. A brief measure for assessing generalized anxiety disorder: the GAD-7. Arch Intern Med. 2006;166(10):1092–7.

34. Johnson SU, Ulvenes PG, Øktedalen T, Hoffart A. Psychometric properties of the general anxiety disorder 7-item (GAD-7) scale in a heterogeneous psychiatric sample. Front Psychol. 2019;10:1713.

35. Prikner O. Vybrané psychometrické charakteristiky škály GAD-7 [Internet]. Masaryk University; 2021. Available from: https://is.muni.cz/th/imwek/

36. R Core Team. R: A language and environment for statistical computing. Vienna, Austria: R Foundation for Statistical Computing; 2021.

37. Bakdash JZ, Marusich LR. rmcorr: Repeated Measures Correlation. R package version 0.4.6 [Internet]. 2022. Available from: https://cran.r-project.org/package=rmcorr

38. Kuznetsova A, Brockhoff PB, Christensen RHB. lmerTest Package: Tests in Linear Mixed Effects Models. J Stat Softw [Internet]. 2017;82(13):1–26. Available from: http://www.jstatsoft.org/v82/i13/

39. Pinheiro JC, Bates DM. Mixed-Effects Models in S and S-PLUS [Internet]. New York: Springer-Verlag; 2000. (Statistics and Computing). Available from: http://link.springer.com/10.1007/b98882

40. Gignac GE, Szodorai ET. Effect size guidelines for individual differences researchers. Pers Individ Dif [Internet]. 2016;102:74–8. Available from: https://www.sciencedirect.com/science/article/pii/S0191886916308194

41. Bazzani A, Bruno S, Frumento P, Cruz-Sanabria F, Turchetti G, Faraguna U. Sleep quality mediates the effect of chronotype on resilience in the time of COVID-19. Chronobiol Int [Internet]. 2021 Jun 3;38(6):883–92. Available from: https://www.tandfonline.com/doi/full/10.1080/07420528.2021.1895199

42. Salfi F, Amicucci G, Corigliano D, Viselli L, D’Atri A, Tempesta D, et al. Two years after lockdown: Longitudinal trajectories of sleep disturbances and mental health over the <scp>COVID</scp>-19 pandemic, and the effects of age, gender and chronotype. J Sleep Res [Internet]. 2022 Nov; Available from: https://onlinelibrary.wiley.com/doi/10.1111/jsr.13767

43. Salehinejad MA, Majidinezhad M, Ghanavati E, Kouestanian S, Vicario CM, Nitsche MA, et al. Negative impact of COVID-19 pandemic on sleep quantitative parameters, quality, and circadian alignment: Implications for health and psychological well-being. EXCLI J. 2020;19:1297–308.

44. Beck F, Léger D, Fressard L, Peretti-Watel P, Verger P. Covid-19 health crisis and lockdown associated with high level of sleep complaints and hypnotic uptake at the population level. J Sleep Res [Internet]. 2021 Feb 28;30(1). Available from: https://onlinelibrary.wiley.com/doi/10.1111/jsr.13119

45. Mandelkorn U, Genzer S, Choshen-Hillel S, Reiter J, Meira e Cruz M, Hochner H, et al. Escalation of sleep disturbances amid the COVID-19 pandemic: a cross-sectional international study. J Clin Sleep Med [Internet]. 2021 Jan;17(1):45–53. Available from: http://jcsm.aasm.org/doi/10.5664/jcsm.8800

46. Pinto J, van Zeller M, Amorim P, Pimentel A, Dantas P, Eusébio E, et al. Sleep quality in times of Covid-19 pandemic. Sleep Med [Internet]. 2020 Oct;74:81–5. Available from: https://linkinghub.elsevier.com/retrieve/pii/S1389945720303130

47. Marelli S, Castelnuovo A, Somma A, Castronovo V, Mombelli S, Bottoni D, et al. Impact of COVID-19 lockdown on sleep quality in university students and administration staff. J Neurol [Internet]. 2021 Jan 11;268(1):8–15. Available from: https://link.springer.com/10.1007/s00415-020-10056-6

48. Casagrande M, Favieri F, Tambelli R, Forte G. The enemy who sealed the world: effects quarantine due to the COVID-19 on sleep quality, anxiety, and psychological distress in the Italian population. Sleep Med [Internet]. 2020 Nov;75:12–20. Available from: https://linkinghub.elsevier.com/retrieve/pii/S1389945720302136

49. Cellini N, Conte F, De Rosa O, Giganti F, Malloggi S, Reyt M, et al. Changes in sleep timing and subjective sleep quality during the COVID-19 lockdown in Italy and Belgium: age, gender and working status as modulating factors. Sleep Med [Internet]. 2021;77:112–9. Available from: https://www.sciencedirect.com/science/article/pii/S138994572030527X

50. Bessot N, Langeard A, Dosseville F, Quarck G, Freret T. Chronotype influence on the effects of <scp>COVID</scp>-19 lockdown on sleep and psychological status in France. J Sleep Res [Internet]. 2023 Feb 20; Available from: https://onlinelibrary.wiley.com/doi/10.1111/jsr.13864

51. Fabbian F, Zucchi B, De Giorgi A, Tiseo R, Boari B, Salmi R, et al. Chronotype, gender and general health. Chronobiol Int [Internet]. 2016 Aug 8;33(7):863–82. Available from: https://www.tandfonline.com/doi/full/10.1080/07420528.2016.1176927

52. Rique GLN, Fernandes Filho GMC, Ferreira ADC, de Sousa-Muñoz RL. Relationship between chronotype and quality of sleep in medical students at the Federal University of Paraiba, Brazil. Sleep Sci [Internet]. 2014 Jun;7(2):96–102. Available from: http://linkinghub.elsevier.com/retrieve/pii/S1984006314000315

53. Merikanto I, Kronholm E, Peltonen M, Laatikainen T, Lahti T, Partonen T. Relation of Chronotype to Sleep Complaints in the General Finnish Population. Chronobiol Int [Internet]. 2012 Apr 6;29(3):311–7. Available from: http://www.tandfonline.com/doi/full/10.3109/07420528.2012.655870

54. Staller N, Randler C. Changes in sleep schedule and chronotype due to COVID-19 restrictions and home office. Somnologie Schlafforsch und Schlafmedizin = Somnology sleep Res sleep Med. 2021;25(2):131–7.

55. Salfi F, Lauriola M, Amicucci G, Corigliano D, Viselli L, Tempesta D, et al. Gender-related time course of sleep disturbances and psychological symptoms during the COVID-19 lockdown: A longitudinal study on the Italian population. Neurobiol Stress [Internet]. 2020 Nov;13:100259. Available from: https://linkinghub.elsevier.com/retrieve/pii/S2352289520300497

56. Neculicioiu VS, Colosi IA, Costache C, Sevastre-Berghian A, Clichici S. Time to Sleep?&mdash;A Review of the Impact of the COVID-19 Pandemic on Sleep and Mental Health. Vol. 19, International Journal of Environmental Research and Public Health. 2022.

57. Kocevska D, Blanken TF, Van Someren EJW, Rösler L. Sleep quality during the COVID-19 pandemic: not one size fits all. Sleep Med [Internet]. 2020 Dec;76:86–8. Available from: https://linkinghub.elsevier.com/retrieve/pii/S1389945720304408

58. Meaklim H, Junge MF, Varma P, Finck WA, Jackson ML. Pre-existing and post-pandemic insomnia symptoms are associated with high levels of stress, anxiety, and depression globally during the COVID-19 pandemic. J Clin Sleep Med [Internet]. 2021 Oct;17(10):2085–97. Available from: http://jcsm.aasm.org/doi/10.5664/jcsm.9354

59. Jahrami H, BaHammam AS, Bragazzi NL, Saif Z, Faris M, Vitiello M V. Sleep problems during the COVID-19 pandemic by population: a systematic review and meta-analysis. J Clin Sleep Med [Internet]. 2021 Feb;17(2):299–313. Available from: http://jcsm.aasm.org/doi/10.5664/jcsm.8930

60. Jahrami HA, Alhaj OA, Humood AM, Alenezi AF, Fekih-Romdhane F, AlRasheed MM, et al. Sleep disturbances during the COVID-19 pandemic: A systematic review, meta-analysis, and meta-regression. Sleep Med Rev [Internet]. 2022 Apr;62:101591. Available from: https://linkinghub.elsevier.com/retrieve/pii/S1087079222000041

61. Madden PAF, Heath AC, Rosenthal NE, Martin NG. Seasonal changes in mood and behavior: the role of genetic factors. Arch Gen Psychiatry. 1996;53(1):47–55.

62. Harmatz MG, Well AD, Overtree CE, Kawamura KY, Rosal M, Ockene IS. Seasonal variation of depression and other moods: a longitudinal approach. J Biol Rhythms. 2000;15(4):344–50.

63. Winthorst WH, Bos EH, Roest AM, de Jonge P. Seasonality of mood and affect in a large general population sample. PLoS One [Internet]. 2020 Sep 14;15(9):e0239033. Available from: 10.1371/journal.pone.0239033

64. João KADR, Jesus SN de, Carmo C, Pinto P. The impact of sleep quality on the mental health of a non-clinical population. Sleep Med [Internet]. 2018 Jun;46:69–73. Available from: https://www.sciencedirect.com/science/article/pii/S1389945718300698

65. Alvaro PK, Roberts RM, Harris JK. A Systematic Review Assessing Bidirectionality between Sleep Disturbances, Anxiety, and Depression. Sleep [Internet]. 2013 Jul 1;36(7):1059–68. Available from: 10.5665/sleep.2810

66. Afonso P, Fonseca M, Teodoro T. Evaluation of anxiety, depression and sleep quality in full-time teleworkers. J Public Health (Bangkok) [Internet]. 2022 Dec 1;44(4):797–804. Available from: 10.1093/pubmed/fdab164

67. O’Regan D, Jackson ML, Young AH, Rosenzweig I. Understanding the Impact of the COVID-19 Pandemic, Lockdowns and Social Isolation on Sleep Quality. Nat Sci Sleep. 2021;13:2053–64.

68. Altena E, Baglioni C, Espie CA, Ellis J, Gavriloff D, Holzinger B, et al. Dealing with sleep problems during home confinement due to the COVID-19 outbreak: Practical recommendations from a task force of the European CBT-I Academy. J Sleep Res [Internet]. 2020 Aug 4;29(4). Available from: https://onlinelibrary.wiley.com/doi/10.1111/jsr.13052

